# A Global Systematic Review and Meta-Analysis of Sex Disparities in Outcomes in Patients with ST-segment Elevation Myocardial Infarction

**DOI:** 10.64898/2026.01.02.26343368

**Authors:** Hao-Min Cheo, Felix Lee, Arthur Lee, Xavier Ong, Lin-Pin Koh, Mark Y Chan, Ian Wee, Jeanne Ong, Ching-Hui Sia

## Abstract

**Background:** Sex disparities in the delivery of care and in-hospital outcomes following ST-segment elevation myocardial infarction (STEMI) remain a global concern. We performed an updated global meta-analysis comparing clinical outcomes between males and females with STEMI.

**Methods:** This study was conducted according to PRISMA guidelines (PROSPERO CRD42024612510). We searched major electronic databases from 1^st^ January 2000 to 7^th^ November 2024. Pairwise meta-analysis was performed on outcomes related to time-to-therapy, in-hospital outcomes, and optimal therapy, with subgroup analyses of geographic regions. Meta-regression was performed to evaluate heterogeneity.

**Results:** A total of 69 studies, involving 891,585 patients (262,773 females; 628,401 males) were included. Males were younger (MD -7.387 years), less likely to have diabetes, hypertension, and prior stroke, had significantly lower risk of in-hospital mortality (RR 0.56, 95%CI 0.53 to 0.61, P<0.05) and major bleeding (RR 0.59, 95%CI 0.50 to 0.71, P<0.05). Males had significantly shorter time to first medical contact (MD - 32.42mins), door-to-balloon (MD -6.17mins) and door-to-needle (MD -5.53mins) times, more likely to undergo PCI (OR 1.34, 95%CI 1.20 to 1.48, P<0.05), receive P2Y12 inhibitors (OR 1,52, 95%CI 1.04 to 2.23, P=0.03), GP IIb/IIIa inhibitors (OR 1.30, 95%CI 1.14 to 1.49, P<0.05), and ACE inhibitors (OR 1.41, 95%CI 0.92 to 2.17, P=0.11). No differences were observed for aspirin and beta-blocker use. Meta-regression showed female sex (β = 1.40) and DM (β = 0.78) were positively correlated with mortality.

**Conclusion:** Females with STEMI experience longer treatment delays, worse in-hospital outcomes, and suboptimal care. There is an urgent need to address sex disparities in STEMI care.

## Introduction

Globally, ischemic heart disease is the leading cause of death [1]. Over the past 2-decades, advancements in care have led to reduced morbidity and mortality [2–7]. However, sex disparities in the quality of in-hospital care and clinical outcomes continue to persist. Women often experience higher mortality rates, attributed to different demographic and risk factors but importantly also differences in treatment approaches, and delays in receiving medical contact [8]. Despite the widespread implementation of standardised care systems for STEMI worldwide, it remains unclear whether these sex gaps in outcomes have been fully addressed or continue to exist on a global scale. Therefore, understanding and addressing these differences is critical to ensuring equitable care for all patients.

In 2021, Shah et al. [9] conducted a meta-analysis investigating sex disparities in in-hospital care and outcomes for STEMI patients globally. Their study revealed that female patients experienced significantly longer delays to first medical contact and door-to-balloon time, as well as higher rates of in-hospital mortality, repeat myocardial infarction, and major bleeding compared to male patients. However, treatment of STEMI has advanced significantly in recent years, with developments of prehospital thrombolytics [10] to improve the timeliness of STEMI management and mobile health (mHealth) applications to enhance coordination of care [10, 11]. These advancements necessitate an updated analysis to capture their potential impact on sex disparities in STEMI outcomes. With newer studies investigating sex-specific outcomes of STEMI published, this study builds upon the findings of Shah et al., incorporating recent research to provide a comprehensive, up-to-date understanding of sex-based differences in care delays, treatment strategies, and outcomes for patients with STEMI.

## Methods

This study was performed in strict accordance to the *Cochrane Handbook for Systematic Reviews of Interventions* [12], and the Preferred Reporting Items for Systematic Reviews and Meta-Analyses (PRISMA) statement guidelines [13]. The study’s protocol was prospectively registered with PROSPERO (CRD42024612510).

## Search process

An electronic search was comprehensively performed on 7^th^ November 2024 in the following major databases: MEDLINE (via PubMed), the Cochrane databases including CENTRAL (Cochrane Central Register of Controlled Trials), Embase and Scopus, to identify all published studies and abstracts comparing outcomes between male and female patients who were admitted into a hospital for ST-elevated myocardial infarction (STEMI). In consult with a research librarian, a repetitive and exhaustive combination of the following Medical Subject Headings (MeSH) search terms were used: (((myocardial infarction[MeSH Terms]) OR (STEMI[MeSH Terms])) AND (sex OR gender)) and (mortality[MeSH Terms]). A manual search of the reference lists of relevant studies was also done to identify additional relevant studies.

## Inclusion and exclusion criteria

Both randomized controlled trials (RCTs) and non-RCTs were included if comparative outcomes were reported for female and male patients admitted into a hospital for STEMI. The studies must have reported sex specific baseline data and outcomes of interest. Given the lack of an available translator, non-English studies were excluded. Other studies of the following designs were also excluded: surveys, trial protocols, review articles, and opinion pieces. Abstracts with no extractable data were also excluded.

## Selection of studies and outcomes of interest

The selection of studies was performed in two stages by two reviewers independently (Figure 1). Studies were first screened for potential inclusion by their titles and abstracts, and the full texts of these preliminarily included studies were then reviewed in their entirety to confirm their inclusion. The senior authors served as the arbiter to resolve differences of opinion regarding the studies’ eligibility through discussion. In line with the aim of this study, the primary outcomes of interest can be broadly categorized into 1) time to treatment: time to first medical contact (FMC), door-to-balloon (DTB), door-to-needle (DTN); and 2) in-hospital outcomes: mortality, repeat MI, stroke, major bleeding. Secondary outcomes include whether patients received optimal therapy, which includes percutaneous coronary intervention (PCI), aspirin, P2Y12 inhibitors, GP IIa/IIIb inhibitors, beta-blockers, and angiotensin converting enzyme (ACE) inhibitors. In addition, the following data were obtained from each study: first author, year, study design, age, presence of comorbidities including diabetes mellitus (DM), hypertension (HTN), smoking status, prior MI, prior stroke, and cardiogenic shock. These data were extracted using a prespecified proforma sheet.

**Figure 1:**
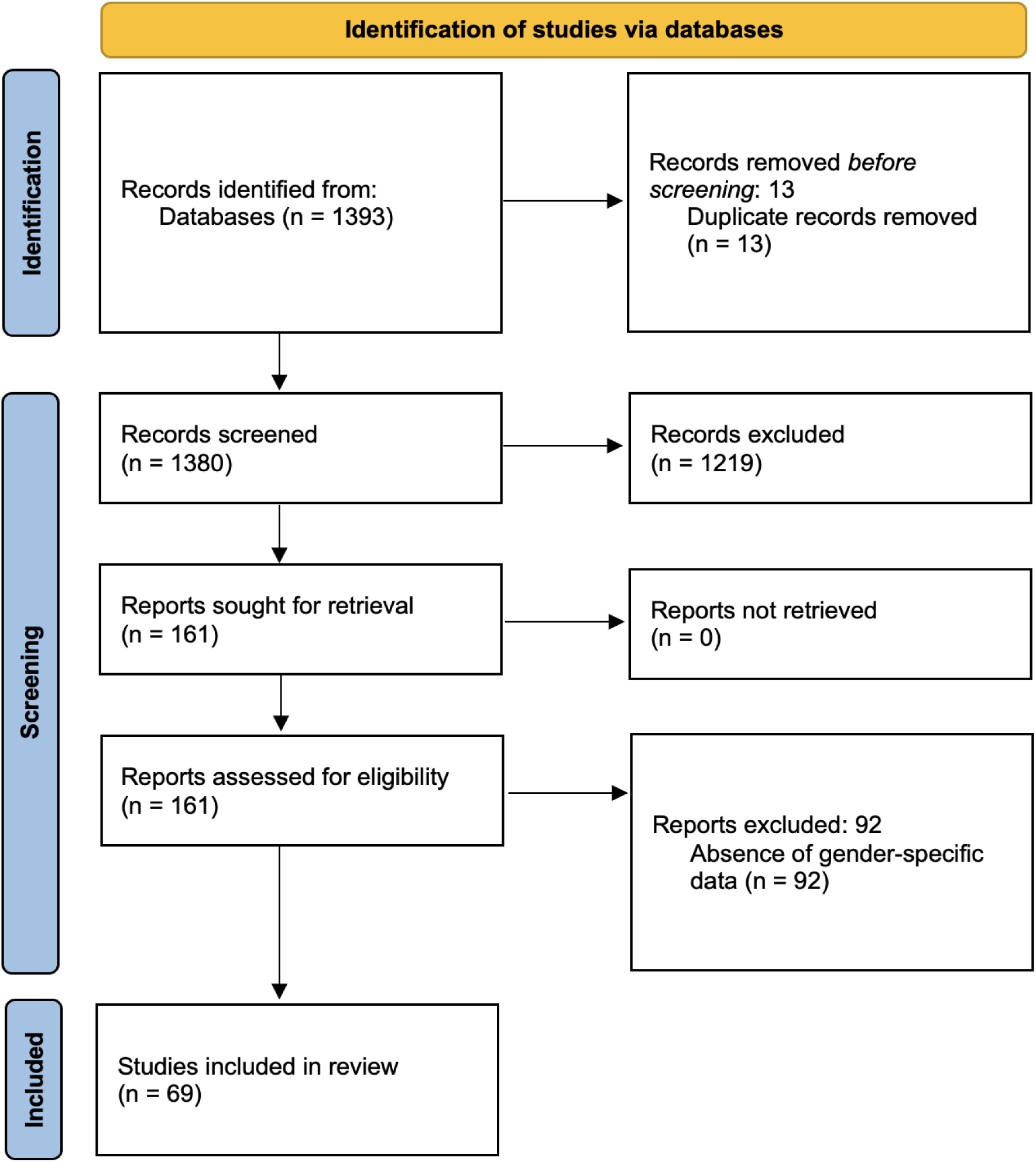
PRISMA flow diagram depicting study selection process

## Statistical analysis

All statistical analyses were performed on STATA (StataCorp. 2025. Stata Statistical Software: Release 18.5. College Station, TX: StataCorp LLC). Pooled weighted mean differences or standardized mean differences were used as the summary statistics for continuous variables, while either the odds ratio (OR) or risk ratio (RR) were employed for dichotomous variables as deemed fit. The I^2^ value was computed to estimate statistical heterogeneity, and a random-effects model was chosen when the value was greater than 50%. The results were reported with 95% confidence intervals (CIs), and a P-value of less than 0.05 was regarded as statistically significant. Variables reported as median (range) were converted to the respective mean and standard deviation using the methods described by Hozo et al. [14] and Wanet et al. [15]. Pre-specified subgroup analysis was performed for all outcomes of interest based on the geographic region of the study, categorized into Asia, Australia, Europe, Middle East and North America. Mixed-effects meta-regression was performed to explore for sources of heterogeneity in the outcomes of interest [16].

## Assessment of Bias

The Cochrane Risk of Bias in Non-randomized Studies – of Exposure (ROBINS-E) tool was utilized to evaluate the risk of bias for observation studies based on seven domains: confounding, measurement of exposure, selection bias, post-exposure interventions, missing data, measurement of outcome and selection of reported results [17]. For outcomes with greater than 10 studies, publication bias was assessed using either funnel plots or the Egger regression test [11].

## Results

### Systematic search

The initial search across all databases yielded 1,393 citations of which 1381 remained after removal of duplicates. After title and abstract screening, 162 articles remained and were subsequently reviewed in their entirety. 69 articles were finally included after full text screening. Reasons for exclusion at each stage are listed in Figure 1.

### Study characteristics

In total, 891,585 patients were included, of whom 262,773 were female and 628,401 were male. The mean age of the male group was significantly younger than the female group (MD -7.387 years, 95%CI -8.053 to -6.720) suggesting that the age of presentation of STEMI was younger for males. When pooling the various comorbidities, females were more likely to have DM (27.8% vs 20.3%), HTN (60.0% vs 46.6%), prior stroke (6.7% vs 4.6%), and present with cardiogenic shock (12.2% vs 10.2%) than males. Males were more likely to be smokers (48.6% vs 26.0%) and have prior MI (13.4% vs 11.2%). The detailed baseline characteristics can be found in Table 1 (Supplementary File 1).

### Study quality

Publication bias as assessed based on funnel plot asymmetry, did not reveal any significant publication for all outcomes of interest. There was overall low risk of bias across the studies as assessed by ROBINS-E tool (Supplementary Table 1).

### Primary outcomes

#### Time to therapy

Males had a significantly shorter FMC (MD -32.42mins, 95%CI -40.84 to - 24.01mins, P<0.05) as compared to females (Figure 2). Subgroup analysis showed consistent findings across all 5 geographical regions. Door-to-balloon times were also significantly shorter in males as compared to females (MD -6.17mins, 95%CI -9.09 to - 3.26mins, P<0.05) (Figure 3). Subgroup analysis revealed that this finding was consistent for studies in Australia, Europe and North America, however there was no statistically significant subgroup difference (P=0.85). Lastly, males had a significantly shorter DTN time as compared to females (MD -5.53mins, 95%CI -10.16 to -0.90mins, P=0.02) (Figure 4). Subgroup analysis was not performed given limited data.

**Figure 2:**
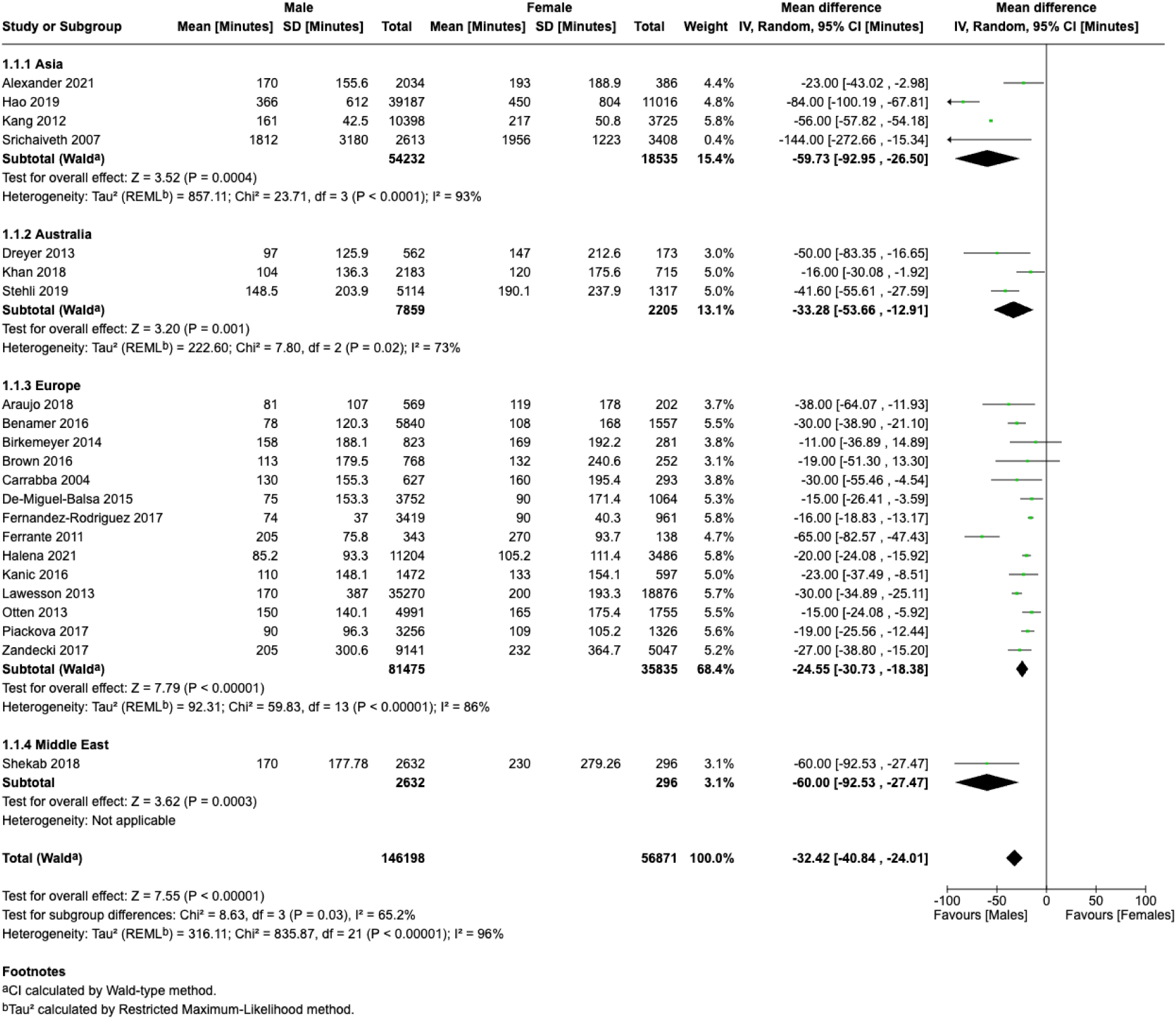
Time to First Medical Contact

**Figure 3:**
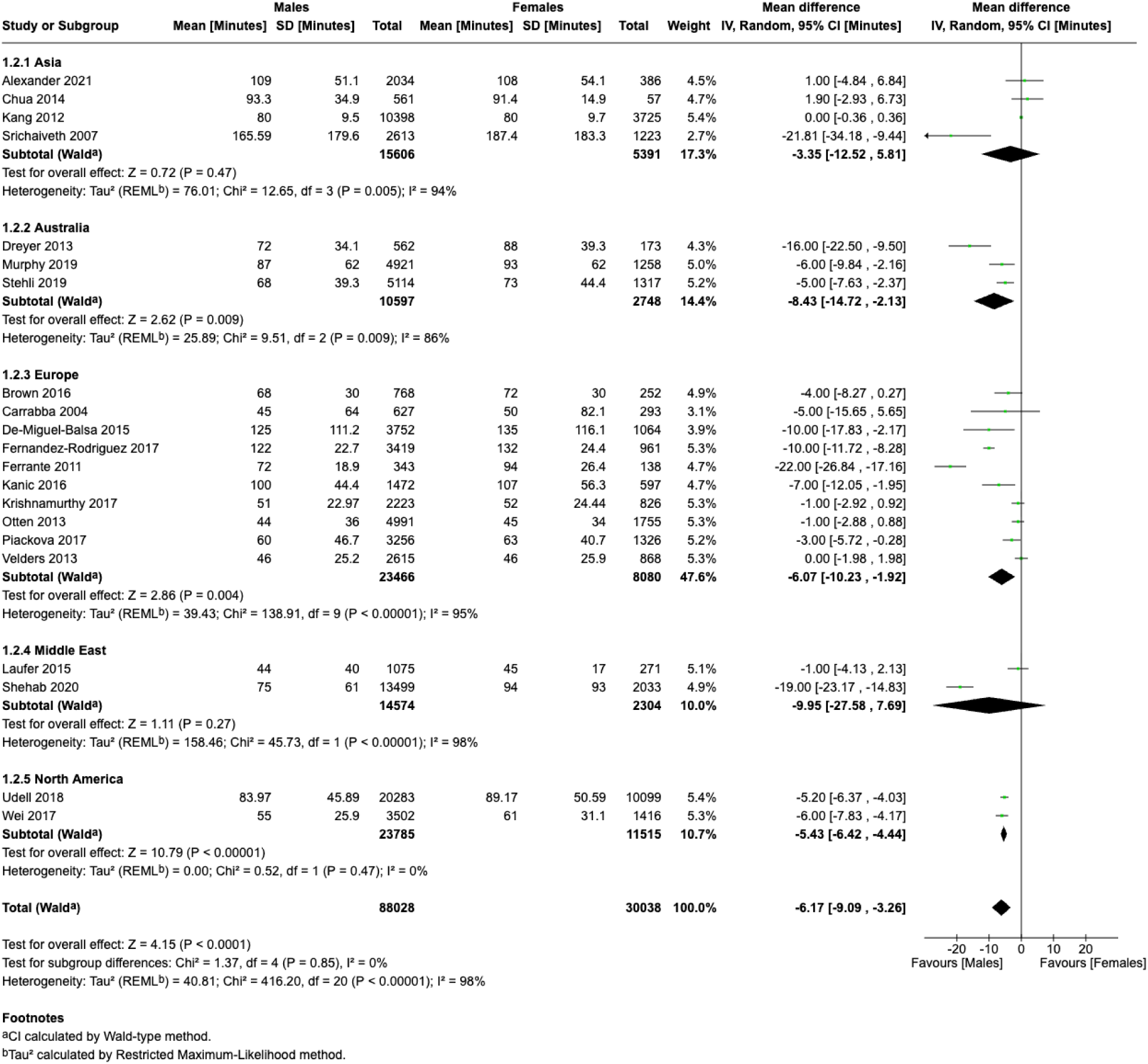
Door-to-Balloon Time

**Figure 4:**
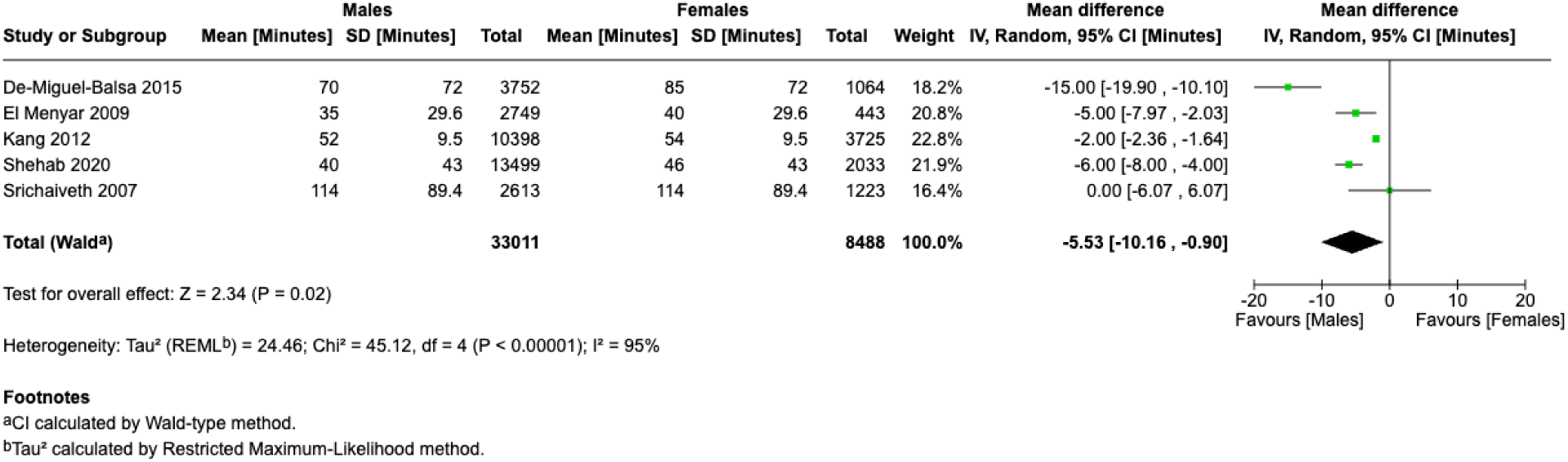
Door-to-Needle

#### In-hospital outcomes

Males had a significantly lower risk of in-hospital mortality as compared to females (RR 0.56, 95%CI 0.53 to 0.61, P<0.05) (Figure 5). Subgroup analysis showed consistent findings for all geographic regions with significant subgroup differences (P=0.00003). Males also had a significantly lower risk of bleeding as compared to females (RR 0.59, 95%CI 0.50 to 0.71, P<0.05), with no significant subgroup differences in geographic regions (Figure 6). Males also had a significantly lower risk of repeat MI (RR 0.79, 95%CI 0.72 to 0.87, P<0.05) (Supplementary Figure 1) and stroke (RR 0.56, 95%CI 0.44 to 0.72, P<0.05) (Supplementary Figure 2) as compared to females. There were significant subgroup differences.

**Figure 5:**
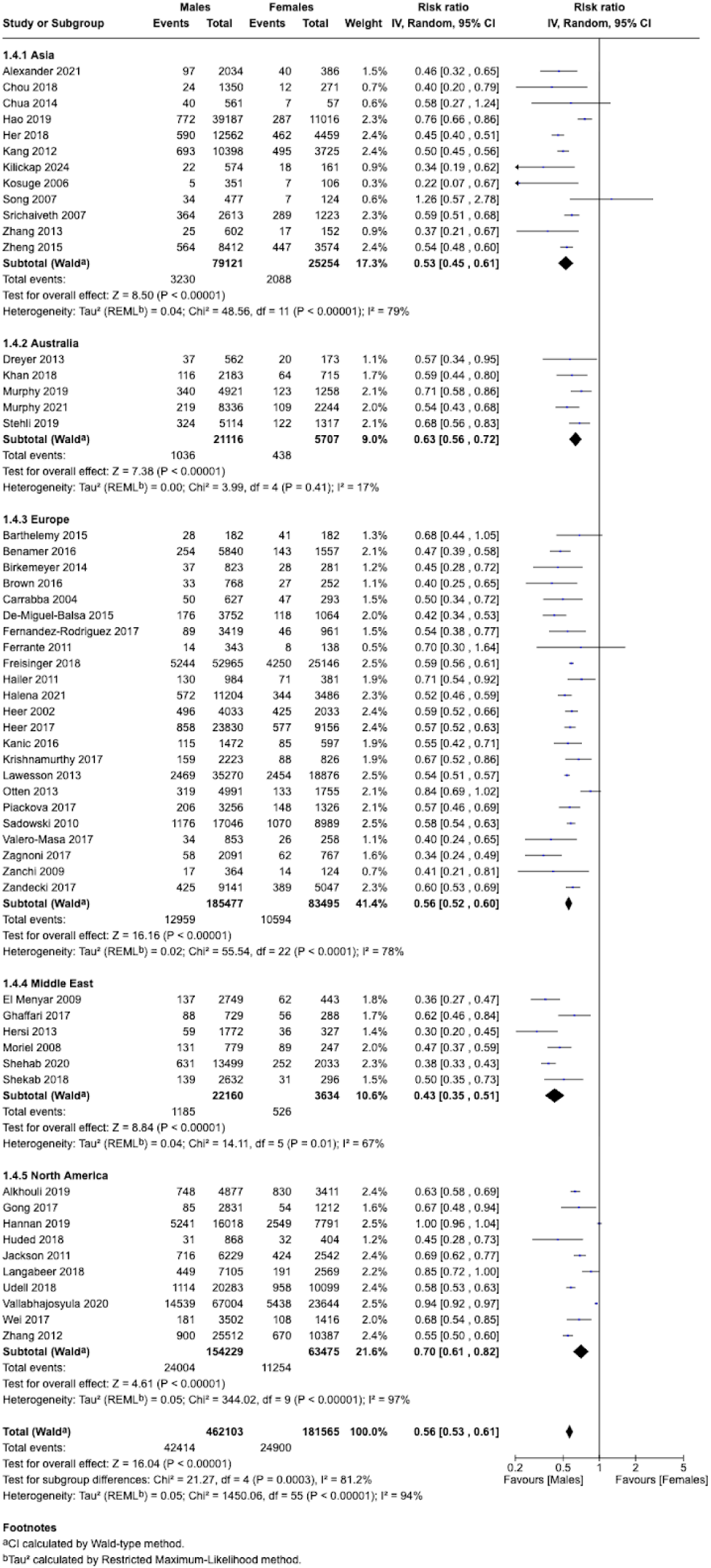
In-Hospital Mortality

**Figure 6:**
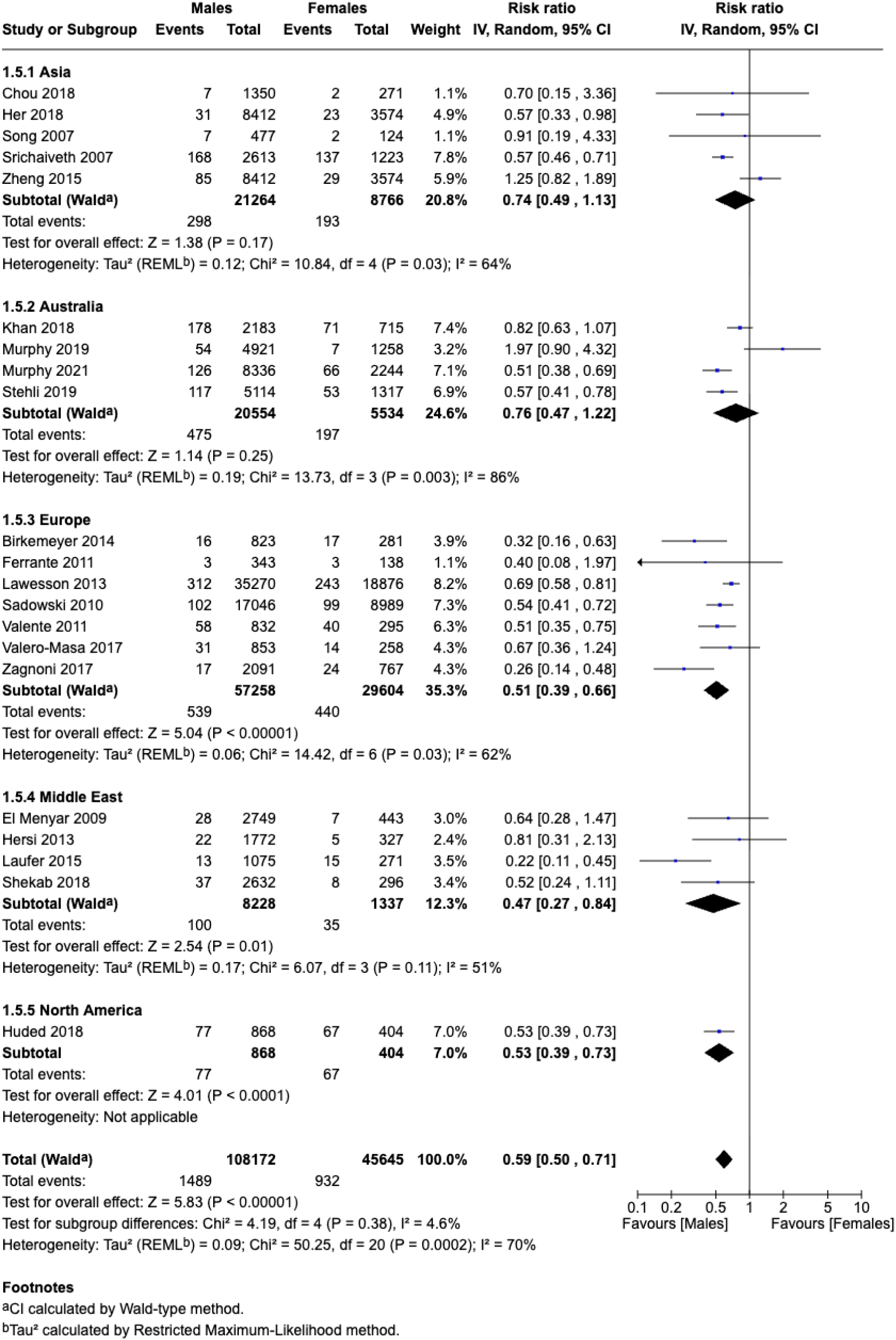
Major Bleeding

### Secondary outcomes

#### Optimal therapy

Males were more likely to undergo PCI as compared to females (OR 1.34, 95%CI 1.20 to 1.48, P<0.05) (Supplementary Figure 3), and this was observed for studies conducted in all regions except the Middle East, where the difference was not statistically significant. There was no significant subgroup difference. In terms of pharmacotherapy, there was no difference in terms of the odds of receiving aspirin between both sexes (OR 1.03, 95%CI 0.73 to 1.46, P=0.85) (Supplementary Figure 4). Only studies conducted in Australia showed that males were more likely to receive aspirin as compared to females, however the subgroup difference was not statistically significant (P=0.50). In addition, there was no significant differences in the odds of receiving ACE inhibitors between both genders (OR 1.41, 95%CI 0.92 to 2.17, P=0.11) (Supplementary Figure 5). However, the odds of receiving P2Y12 inhibitors (OR 1.52, 95%CI 1.04 to 2.23, P=0.03) (Supplementary Figure 6) was higher in males. Subgroup analysis revealed significant differences between geographic regions (P=0.007), where the odds of receiving P2Y12 inhibitors amongst males were higher in Australia and Europe. The odds of receiving GP IIb/IIIa inhibitors were higher in males as compared to females (OR 1.30, 95%CI 1.14 to 1.49, P<0.05) (Supplementary Figure 7), with no significant subgroup differences. Lastly, males were also more likely to receive beta blockers as compared to females (OR 1.25, 95%CI 1.04 to 1.52, P=0.02) (Supplementary Figure 8), with no significant subgroup differences.

### Meta-regression

Meta-regression was conducted on the pooled proportion of mortality, which showed that female sex (β = 1.40) and DM (β = 0.78) were positively correlated with mortality (Supplementary Figures 9 and 10). The other variables in the model including age, smoking status and aspirin were not significant. Meta-regression models for repeat MI and major bleeding did not show any significant variables.

## Discussion

To our knowledge, this meta-analysis provides the most contemporary and comprehensive global assessment of sex-based disparities in the management and outcomes of STEMI. Consistent with the previous meta-analysis in 2021 [9], females continue to experience longer delays to care, poorer in-hospital outcomes and a lower likelihood of receiving optimal therapy. These disparities persist despite overall advancements in STEMI care over the past two decades. Compared to the previous meta-analysis by Shah et al. [9], we have provided an updated evidence base, regional subgroup analyses, and novel meta-regression findings.

Globally, the management of STEMI has seen substantial progress, in part due to international efforts like the Global Heart Attack Treatment Initiative (GHATI) program led by the American College of Cardiology (ACC) [18]. Over the initial two-year study period (2020-2021) of the GHATI program, sustained improvements were observed across both clinical and system-level measures. These included reductions in the composite outcome of cardiogenic shock at presentation, cardiac arrest pre- and post-intervention, reduced final ejection fraction (< 40%), and improved survival at discharge. Consistent adherence to guideline-directed medical therapies and shorter transportation times further reflect the positive influence of evidence-based guidelines, even in settings with limited resources. [19] These advancements highlight the importance of standardized STEMI care protocols in reducing overall mortality and morbidity. Nevertheless, despite improvements in STEMI care, our findings suggest that sex-based disparities in STEMI care may not have improved at the same pace.

Overall, the global findings demonstrated consistent sex disparities in outcomes across all regions. Females experienced significantly longer delays to care compared to males, particularly in time to FMC, with a mean delay of 37.8 minutes. While the differences in DTB time across sex were less noticeable, they remained statistically significant with mean delays nearing seven minutes. Regional delays in DTB exceeding five mins were observed in Australia, Europe, North America, and the Middle East. Furthermore, females consistently exhibited higher rates of in-hospital mortality across all regions, as well as higher rates of major bleeding in Asia, Europe, North America, and the Middle East. These findings highlight the pervasive nature of sex-based inequities in acute care and outcomes for female patients with STEMI, regardless of geographical differences. Despite so, we can still observe that many geographical regions do not have published data on sex disparities in outcomes of patients with STEMI (grey areas in Supplementary Figure 11), and that is a gap that still needs addressing.

Our study found that women experienced longer delays than men at nearly every step from the onset of symptoms to receiving revascularization treatment. While earlier research pointed to patient delay—especially women waiting longer before seeking help—as the main cause [20, 21], our findings confirm that women do wait longer for emergency services (EMS) after symptoms begin. This delay may be due to women misjudging their symptoms as non-cardiac or delaying care because of other reasons such as caregiving responsibilities [22]. Other contributing factors may include lower socioeconomic status, which is linked to reduced awareness of heart disease risk, and broader sex-related issues [22,23]. Still, our results highlight that the most significant delays in women with STEMI came from the healthcare system itself—both before reaching the hospital and within it. Women with heart attacks often have non-specific symptoms like nausea, radiating pain, or shortness of breath, which may not be recognized as signs of STEMI [22]. Healthcare providers may be less likely to identify these symptoms as serious in women compared to men, potentially leading to incorrect triage and delays in care [22–24]. Notably, even women with classic symptoms have been shown to face delays, pointing to a potential bias in how healthcare workers perceive heart attack risk in women [23].

Women with STEMI experience poorer outcomes including higher risks of mortality and bleeding, and this could be due to a myriad of reasons. Consistent with our findings they tend to present at an older age with increased comorbidities, and consequently poorer cardiovascular risk profiles [25,26]. Beyond baseline differences in cardiovascular risk factors, delays in care and differences in STEMI management can also contribute to poorer outcomes. Data from the National Cardiovascular Data Registry’s Acute Coronary Treatment and Intervention Outcomes Network Registry—Get With The Guidelines—showed that between 2008 and 2014, women continued to have longer contact-to-device times than men, resulting in higher mortality rates in both sexes [27]. This finding aligns with the VIRGO study, which observed that younger patients who did not receive timely reperfusion—especially those transferred for PCI—had higher short- and long-term mortality rates compared to those who were treated within the recommended time frames [28]. These highlight the importance of timely treatment, suggesting that STEMI treatment disparities and mortality in women can in fact be overcome using standardized STEMI protocols and systems. The higher risk of bleeding amongst females could arise from inappropriate dosing of antithrombotic agents, given sex differences in pharmacodynamics and pharmacokinetics [29]. This may indicate that current pharmacotherapy guidelines fail to account for sex-specific differences in treatment needs. Since guidelines are typically developed from clinical trial data—and women are often underrepresented in these trials [30]—the resulting recommendations could lack generalisability and may not fully reflect the needs of female patients.

Sex-based differences in the underlying pathophysiology of STEMI may contribute to variations in how patients receive and respond to guideline-directed therapy. For instance, women are less likely than men to have a clearly identifiable culprit lesion during angiography and are more often diagnosed with non-obstructive coronary artery disease [31]. Myocardial infarction with non-obstructive coronary arteries (MINOCA) is more prevalent in women, and can result from several mechanisms, including plaque rupture or ulceration, coronary vasospasm, embolism, spontaneous coronary artery dissection, and Takotsubo cardiomyopathy [32]. Despite its clinical significance, there are currently no standardized national guidelines for diagnosing and managing MINOCA. The VIRGO study found that about one in eight young women with acute myocardial infarction did not meet the universal diagnostic criteria, prompting researchers to propose a new classification system aimed at better identifying patient subtypes and guiding treatment [28]. Future studies should integrate imaging or registry-level data to better define these phenotypes.

As compared to Shah et al.’s study [9], females in our study did not experience significantly higher rates of repeat MI as compared to males. This was despite our study also highlighting significantly lower levels of P2Y12 inhibitor usage and lower rates of primary PCI in females. This could be partially due to biological differences between males and females, with females being shown to have complete ST-resolution and smaller infarct sizes as compared to males [33]. Besides this, the age difference in male and females in Shah et al. [9] study was greater than in our study. This could potentially explain the difference in findings, as increased age has been suggested to increase likelihood of repeat MI [34]. Certain findings in Shah’s et al.’s study has been echoed by our study, namely the increased risk of in-hospital mortality and major bleeding for female patients as compared to male patients, alongside significantly lower times to FMC and significantly door-to-balloon times in males as compared to females. These findings suggest sex-based disparities in STEMI care and potential areas for targeted improvement in the management of female patients.

Nonetheless, the findings of our study must be interpreted in the context of known limitations. The inclusion of retrospective studies would raise issues of bias and heterogeneity, especially given variations in patient demographics and healthcare systems across different geographic regions. To address this, we performed subgroup analysis based on geographical regions to determine differences that could have explained the heterogeneity. Furthermore, we conducted a thorough risk of bias assessment and formal analysis of publication bias, both of which did not reveal any significant areas of bias. Lastly, we performed meta-regression analysis as means of identifying potential sources of heterogeneity, which were consistent with our initial meta-analysis findings showing that female sex was significantly associated with mortality. Some subgroup analyses may be underpowered (Figures 2, 3, and 6), as some regions included only one or two studies. Next, our study was unable to elucidate possible reasons for some of these findings. This, for instance, can include sociocultural, financial and structural barriers to assessing proper medical care. Future population-based research should consider evaluating these factors, so targeted interventions can be designed to overcome these issues and improve outcomes for women with STEMI.

## Conclusion

In summary, this global meta-analysis shows that women experience significantly higher rates of in-hospital mortality and major bleeding, and this could be a result of poorer cardiovascular risk profiles, delays in receiving care, and less-than-optimal treatment. Despite improvements in STEMI care, this has not necessarily translated to both sexes and more should be done to address these gaps.

## Data Availability

The data that support the findings of this study are openly available in ClinicalTrials.gov.

## References

1. World Health Organization. The top 10 causes of death. Available from: https://www.who.int/news-room/fact-sheets/detail/the-top-10-causes-of-death (accessed 15 Apr 2025).

2. Wong GC, Welsford M, Ainsworth C, Abuzeid W, Fordyce CB, Greene J, et al. 2019 Canadian Cardiovascular Society/Canadian Association of Interventional Cardiology Guidelines on the Acute Management of ST-Elevation Myocardial Infarction: Focused Update on Regionalization and Reperfusion. Can J Cardiol 2019;35(2):107–132. 10.1016/j.cjca.2018.11.031

3. Ibanez B, James S, Agewall S, Antunes MJ, Bucciarelli-Ducci C, Bueno H, et al. 2017 ESC Guidelines for the management of acute myocardial infarction in patients presenting with ST-segment elevation. Eur Heart J 2017;39(2):119–177. 10.1093/eurheartj/ehx393

4. Levine GN, Bates ER, Blankenship JC, Bailey SR, Bittl JA, Cercek B, et al. 2015 ACC/AHA/SCAI focused update on primary percutaneous coronary intervention for patients with ST-elevation myocardial Infarction: An update of the 2011 ACCF/AHA/SCAI guideline for percutaneous coronary intervention and the 2013 ACCF/AHA guideline for the management of ST-elevation myocardial infarction: A report of the American College of Cardiology/American Heart Association Task Force on Clinical Practice Guidelines and the Society For Cardiovascular Angiography and Interventions. Catheter Cardiovasc Interv 2016;87(6):1001–1019. 10.1002/ccd.26325

5. Jacobs AK, Antman EM, Faxon DP, Gregory T, & Solis P. Development of Systems of Care for ST-Elevation Myocardial Infarction Patients. Circulation 2007;116(2):217–230. 10.1161/circulationaha.107.184043

6. Chew DP, Scott IA, Cullen L, French JK, Briffa TG, Tideman PA, et al. National Heart Foundation of Australia and Cardiac Society of Australia and New Zealand: Australian clinical guidelines for the management of acute coronary syndromes 2016. Med J Aust 2016;205(3):128–133. 10.5694/mja16.00368

7. Langabeer JR, DelliFraine J, Fowler R, Jollis JG, Stuart L, Segrest W, et al. Emergency Medical Services as a Strategy for Improving ST-Elevation Myocardial Infarction System Treatment Times. J Emerg Med 2014;46(3):355–362. 10.1016/j.jemermed.2013.08.112

8. Kilickap M, Erol MK, Kayikcioglu M, Kocayigit I, Gitmez M, Can V, et al. (2020). Short and midterm outcomes in patients with acute myocardial infarction: Results of the Nationwide TURKMI Registry. Angiology 2020;72(4):339–347. 10.1177/0003319720975302

9. Shah T, Haimi I, Yang Y, Gaston S, Taoutel R, Mehta S, et al. Meta-Analysis of Gender Disparities in In-hospital Care and Outcomes in Patients with ST-Segment Elevation Myocardial Infarction. Am J Cardiol. 2021;147(15):23–32. 10.1016/j.amjcard.2021.02.015

10. Guy A, Gabers N, Crisfield C, Helmer J, Peterson SC, Ganstal A, et al. Collaborative Heart Attack Management Program (CHAMP): use of prehospital thrombolytics to improve timeliness of STEMI management in British Columbia. BMJ Open Qual 2021;10(4):e001519. 10.1136/bmjoq-2021-001519

11. Abrahim C, Capatina A, Arvind Kalyan-Sundaram, & Lotfi A. Reducing Door-to-Balloon Time Using EMS-initiated App-based Communication. J Soc Cardiovasc Angiogr Interv 2024;3(9):102199–102199. 10.1016/j.jscai.2024.102199

12. Chandler J, Cumpston M, Thomas J, Higgins JPT, Deeks JJ, Clarke MJ. Cochrane handbook for systematic reviews of interventions version 6.3 (updated August 2022). Cochrane, 2022. Available from: https://training.cochrane.org/handbook/archive/v6.3

13. Moher D, Liberati A, Tetzlaff J, Altman DG, PRISMA Group. Preferred Reporting Items for Systematic Reviews and Meta-Analyses the PRISMA statement. PLoS Med 2009;6:e1000097. 10.1371/journal.pmed.1000097

14. Hozo SP, Djulbegovic B, Hozo I. Estimating the mean and variance from the median, range, and the size of a sample. BMC Med Res Methodol 2005;5:13. 10.1186/1471-2288-5-13

15. Wan X, Wang W, Liu J, Tong T. Estimating the sample mean and standard deviation from the sample size, median, range and/or interquartile range. BMC Med Res Methodol 2014;135(14). 10.1186/1471-2288-14-135

16. Harbord RM, & Higgins JPT. Meta-Regression in Stata. Stata J 2008;8(4):493–519. 10.1177/1536867X0800800403

17. Higgins JPT, Morgan RL, Rooney AA, Taylor KW, Thayer KA, Silva RA, et al. A tool to assess risk of bias in non-randomized follow-up studies of exposure effects (ROBINS-E). Environ Int 2024;186. 10.1016/j.envint.2024.108602

18. American College of Cardiology. Transform Acute myocardial infarction care: Global Heart Attack Treatment Initiative. Available from: https://www.acc.org/Membership/Features/2019/08/Transform-Acute-Myocardial-Infarction-Care-Global-Heart-Attack-Treatment-Initiative (accessed 28 Jun 2025)

19. Herrera CJ, Levenson BJ, Natcheva A, Lucca AC, Olsson K, Miki K, et al. Improving STEMI management internationally. JACC Advances 2024;4(1):101438. 10.1016/j.jacadv.2024.101438

20. Stehli J, Martin C, Brennan A, Dinh DT, Lefkovits J, Zaman S. Sex differences persist in time to presentation, revascularization, and mortality in myocardial infarction treated with percutaneous coronary intervention. J Am Heart Assoc. 2019;8:e012161. 10.1161/JAHA.119.012161

21. Benamer H, Bataille S, Tafflet M, Jabre P, Dupas F, Laborne FX, et al. Longer pre-hospital delays and higher mortality in women with STEMI: the e-MUST Registry. EuroIntervention. 2016;12:e542–e549. DOI: 10.4244/EIJV12I5A93.

22. Lawesson SS, Isaksson R-M, Ericsson M, Ängerud K, Thylén I; SymTime Study Group. Gender disparities in first medical contact and delay in ST-elevation myocardial infarction: a prospective multicentre Swedish survey study. BMJ Open. 2018;8:e020211. 10.1136/bmjopen-2017-020211

23. Lichtman JH, Leifheit EC, Safdar B, Bao H, Krumholz HM, Lorenze NP, et al. Sex differences in the presentation and perception of symptoms among young patients with myocardial infarction: evidence from the VIRGO Study (Variation in Recovery: Role of Gender on Outcomes of Young AMI Patients). Circulation. 2018;137:781–790. 10.1161/CIRCULATIONAHA.117.031650

24. Kirchberger I, Heier M, Kuch B, Wende R, Meisinger C. Sex differences in patient-reported symptoms associated with myocardial infarction (from the population-based MONICA/KORA Myocardial Infarction Registry). Am J Cardiol. 2011;107:1585–1589. 10.1016/j.amjcard.2011.01.040

25. Vogel B, Acevedo M, Appelman Y, et al. The *Lancet* women and cardiovascular disease Commission: reducing the global burden by 2030. Lancet. 2021;397:2385–2438. 10.1016/S0140-6736(21)00684-X

26. Yu J, Mehran R, Grinfeld L, Xu K, Nikolsky E, Brodie BR, et al. Sex-based differences in bleeding and long term adverse events after percutaneous coronary intervention for acute myocardial infarction: three year results from the HORIZONS-AMI trial. Catheter Cardiovasc Interv 2015;85:359–68. 10.1002/ccd.25630

27. Roswell RO, Kunkes J, Chen AY, Chiswell K, Iqbal S, Roe MT, et al. Impact of Sex and Contact-to-Device Time on Clinical Outcomes in Acute ST-Segment Elevation Myocardial Infarction-Findings From the National Cardiovascular Data Registry. J Am Heart Assoc 2017;6:e004521. 10.1161/JAHA.116.004521

28. D’Onofrio G, Safdar B, Lichtman JH, Strait KM, Dreyer RP, Geda M, et al. Sex differences in reperfusion in young patients with ST-segment-elevation myocardial infarction: results from the VIRGO study. Circulation 2015;131:1324–32. 10.1161/CIRCULATIONAHA.114.012293

29. Laborante R, Borovac JA, Galli M, Rodolico D, Ciliberti G, Restivo A, et al. Gender-differences in antithrombotic therapy across the spectrum of ischemic heart disease: Time to tackle the Yentl syndrome? Front Cardiovasc Med. 2022;31(9):1009475. 10.3389/fcvm.2022.1009475

30. Jin X, Chandramouli C, Allocco B, Gong E, Lam CSP, Yan LL. Women’s Participation in Cardiovascular Clinical Trials From 2010 to 2017. Circulation. 2020;141(7):540–548. 10.1161/CIRCULATIONAHA.119.043594

31. Wei J, Mehta PK, Grey E, Garberich RF, Hauser R, Merz CNB, et al. Sex-based differences in quality of care and outcomes in a health system using a standardized STEMI protocol. Am Heart J 2017;191:30–36. 10.1016/j.ahj.2017.06.005

32. Agewall S, Beltrame JF, Reynolds HR, Niessner A, Rosano G, Caforio ALP, et al. ESC working group position paper on myocardial infarction with non-obstructive coronary arteries. Eur Heart J 2017;38:143–53. 10.1093/eurheartj/ehw149

33. Ng VG, Mori K, Costa RA, Kish M, Mehran R, Urata H, et al. Impact of gender on infarct size, ST-segment resolution, myocardial blush and clinical outcomes after primary stenting for acute myocardial infarction: Substudy from the EMERALD trial. Intl J Cardiol. 2016 Jan 6;207:269–76. 10.1016/j.ijcard.2016.01.013

34. Al Saleh AS, Alhabib KF, Alsheik-Ali AA, Sulaiman K, Alfaleh H, Alsaif S, et al. Predictors and Impact of In-Hospital Recurrent Myocardial Infarction in Patients With Acute Coronary Syndrome: Findings From Gulf RACE-2. Angiology. 2016 Oct 26;68(6):508–12. 10.1177/0003319716674855

